# Key Histologic Features Distinguish Cytomegalovirus Hepatitis from Acute T-cell Mediated Rejection in Liver Allografts

**DOI:** 10.1101/2025.08.29.25334504

**Authors:** Shunsuke Koga, Yusuf Ozcelik, Liping Wang, Sujani Madabhushi, Kristen M. Stashek, Emma E. Furth, Rashmi Tondon

## Abstract

Cytomegalovirus (CMV) is a major opportunistic infection after liver transplantation and often mimics acute T cell-mediated rejection (TCMR), creating management uncertainty. We conducted a retrospective study to identify practical histologic features that distinguish CMV hepatitis from TCMR in routine practice. We included 10 recipients with CMV hepatitis and 5 with moderate to severe TCMR. Portal inflammation, bile duct injury, venous endotheliitis, lobular microgranulomas, neutrophilic microabscesses, and CMV inclusions were assessed. Clinical data were abstracted from the medical record.

CMV hepatitis was diagnosed earlier after transplantation than TCMR (272 ± 211 vs 549 ± 522 days). Slides were available for 7 CMV and all 5 TCMR biopsies. Histologic findings and their diagnostic performance estimates were as follows: microgranulomas present in 7/7 CMV and 0/5 TCMR biopsies, sensitivity 100% and specificity 100%; bile duct injury minimal to absent in 6/7 CMV and 0/5 TCMR biopsies, sensitivity 86% and specificity 100%; neutrophilic microabscesses in 3/7 CMV and 0/5 TCMR biopsies, sensitivity 42.9% and specificity 100%. Antiviral therapy was administered in 9/10 CMV patients (90%); recurrent CMV viremia occurred in 4/10 (40%) and late chronic rejection in 2/10 (20%), while no CMV viremia occurred in the TCMR group.

In routine practice, a pattern of portal lymphohistiocytic inflammation with lobular microgranulomas and minimal to absent bile duct injury supports CMV hepatitis over TCMR and can guide targeted search for inclusions and CMV PCR, which may help avoid unnecessary intensification of immunosuppression and enable timely antiviral therapy.

## Introduction

Cytomegalovirus (CMV) is one of the most significant opportunistic infections after liver transplantation, particularly in seronegative recipients of seropositive donor grafts (D+/R-) where risk is highest.^1^ Its prevention is stratified by donor–recipient serostatus in contemporary guidelines.^2^ Even with modern prevention, residual risk persists. In a randomized trial of D+/R-liver recipients, 1211month CMV disease occurred in 9% with preemptive therapy versus 19% with universal prophylaxis, while observational data link CMV infection with inferior graft survival.^3, 4^ These data frame CMV as both common and prognostically relevant in liver transplantation and set the stage for accurate diagnosis on biopsy.

Diagnostic separation of CMV hepatitis from acute T cell-mediated rejection (TCMR) is challenging because early clinical and histologic findings overlap, yet initial therapy diverges, with antiviral treatment for CMV and increased immunosuppression for rejection.^5^ The Banff Working Group schema defines acute TCMR by a triad of portal inflammation, bile duct injury, and venous endotheliitis.^6^ Prior studies have described histologic features that raise suspicion for CMV, including lobular microgranulomas and histiocyte-rich inflammation. Presence of classic viral cytopathic inclusions on hematoxylin and eosin (H&E) sections is diagnostic but these inclusions may be sparse, sometimes limited to a single cell.^7^ Recent series emphasize that CMV can mimic acute TCMR at low power; therefore a targeted search for inclusions is required to reach the correct diagnosis.^8^

There is a need for simple morphologic clues that separate CMV hepatitis from acute TCMR in routine sign out. To address this gap, we conducted a retrospective single11center study of liver allograft biopsies to define practical features that guide correct classification and treatment.

## Materials and Methods

### Study design

We conducted a retrospective, single-center study at the Hospital of the University of Pennsylvania (Philadelphia, Pennsylvania) over a 15-year period (7/1/2010 to 6/30/2025). All biopsies had been diagnosed in routine clinical practice by experienced, American Board of Pathology certified anatomic pathologists with expertise in medical liver pathology.^9^

### Case selection

We searched the surgical pathology archives and electronic medical record to identify liver allograft biopsies diagnosed as CMV hepatitis and a comparison group with acute TCMR from the same period. CMV hepatitis required CMV cytopathic inclusions on H&E sections; plasma CMV polymerase chain reaction (PCR) results were recorded when available. Acute TCMR controls were randomly selected biopsies reported as moderate to severe per Banff Working Group criteria and had no detectable plasma CMV by PCR at biopsy.

### Pathology review and data collection

For all patients with available slides, a single pathologist (RT), board-certified in anatomic pathology and specializing in medical liver pathology, re-reviewed H&E sections and recorded histologic features. The variables included portal inflammation, bile duct injury, venous endotheliitis in both portal and central veins, lobular microgranulomas, neutrophilic microabscesses, and CMV inclusions. Clinical information was abstracted from the medical record. Clinical variables included age, sex, indication for transplantation, donor/recipient CMV serostatus, time from transplant to biopsy, CMV prophylaxis, prior or subsequent episodes of CMV or TCMR, treatments at the index biopsy (antiviral and/or antirejection), and follow-up outcomes (incident or recurrent CMV and chronic rejection).

### Statistical analysis

Continuous variables are reported as mean and standard deviation and compared between groups using Welch’s t tests because variances could be unequal in small samples. Categorical variables are compared using two-sided Fisher’s exact tests. For histologic features considered as diagnostic tests, we calculated sensitivity and specificity with exact binomial 95% confidence intervals (CI) using the Clopper–Pearson approach. The significance level was set at 0.05 without adjustment for multiple comparisons given the exploratory intent.

## Results

### Cohort characteristics

We analyzed 10 recipients with CMV hepatitis and 5 with acute TCMR. Baseline demographics, serostatus, and CMV prophylaxis are summarized in **Table 1**. The mean age was 56 years in the CMV group and 50 years in the acute TCMR group; the age difference was not significant (p = 0.29). In the CMV group, primary sclerosing cholangitis was the most frequent indication for transplantation (4/10 patients), followed by non-alcoholic steatohepatitis, autoimmune hepatitis, alcoholic cirrhosis, primary biliary cholangitis, and hepatitis C (1 patient each), with 1 patient classified as unknown. In the acute TCMR group, alcoholic cirrhosis was the most common indication (3/5 patients), followed by autoimmune hepatitis and hepatitis C (1 patient each). CMV prophylaxis had been administered in 9 of 10 CMV patients and in all 5 patients with acute TCMR. Donor and recipient serostatus in the CMV cohort was D+/R− in 5, D+/R+ in 2, D−/R+ in 1, and donor unknown/R− in 2; in the acute TCMR cohort it was D+/R+ in 2, D+/R− in 2, and D−/R+ in 1. Overall, CMV hepatitis was diagnosed earlier after transplantation than acute TCMR (272 ± 211 vs. 549 ± 522 days; p = 0.31). At the time of biopsy, plasma CMV PCR was positive in all tested CMV patients (7 of 7; mean viral load 137,548 IU/mL) and negative in all patients with acute TCMR (0 of 5).

**Table 1.** Demographic and clinical features.

| **Variable** | **CMV hepatitis (n=10)** | **Acute TCMR (n=5)** |
| --- | --- | --- |
| Age, mean (SD), years | 56 (9) | 50 (10) |
| Male sex, n (%) | 7 (70%) | 4 (80%) |
| Time from transplant to biopsy, mean (SD), days | 272 (211) | 549 (522) |
| Donor/recipient serostatus | D+/R– in 5, D+/R+ in 2, D–/R+ in 1, and donor unknown/R– in 2 | D+/R+ in 2, D+/R– in 2, and D–/R+ in 1 |
| CMV prophylaxis | 9/10 (90%) | 5/5 (100%) |
| CMV PCR positive at biopsy | 7/7 (100%) | 0/5 (0%) |
| Diagnosis of TCMR | Present: 1; indeterminate: 4; none: 5 | Present: 5 |

Abbreviations: CMV, cytomegalovirus; TCMR, T-cell mediated rejection; D+/R–, donor-positive/recipient-negative for CMV IgG serostatus; PCR, polymerase chain reaction.

### Histopathology

Slides were available for review in 7 of 10 CMV biopsies and all 5 TCMR biopsies (**Table 2**). In CMV hepatitis, all reviewed patients showed CMV inclusions on H&E; semi-quantitatively, a single inclusion was present in 3 of 7 patients (43%) and more than three inclusions in 4 of 7 (57%). Endotheliitis was present in all 7 patients. The portal infiltrate was lymphohistiocytic with rare to occasional eosinophils (7/7, 100%), and lobular microgranulomas were identified in every patient (7/7, 100%); one patient shows well-defined portal and lobular non-necrotizing granulomas (14%). Bile duct injury was none or minimal in 6 of 7 patients (86%). Neutrophilic microabscesses were present in 3 of 7 (43%). In contrast, acute TCMR biopsies showed no CMV inclusions, microgranulomas, or neutrophilic microabscesses, and instead demonstrated predominantly lymphocytic portal inflammation with prominent bile duct injury (5/5, 100%), and venous endotheliitis (5/5, 100%).

**Table 2:**
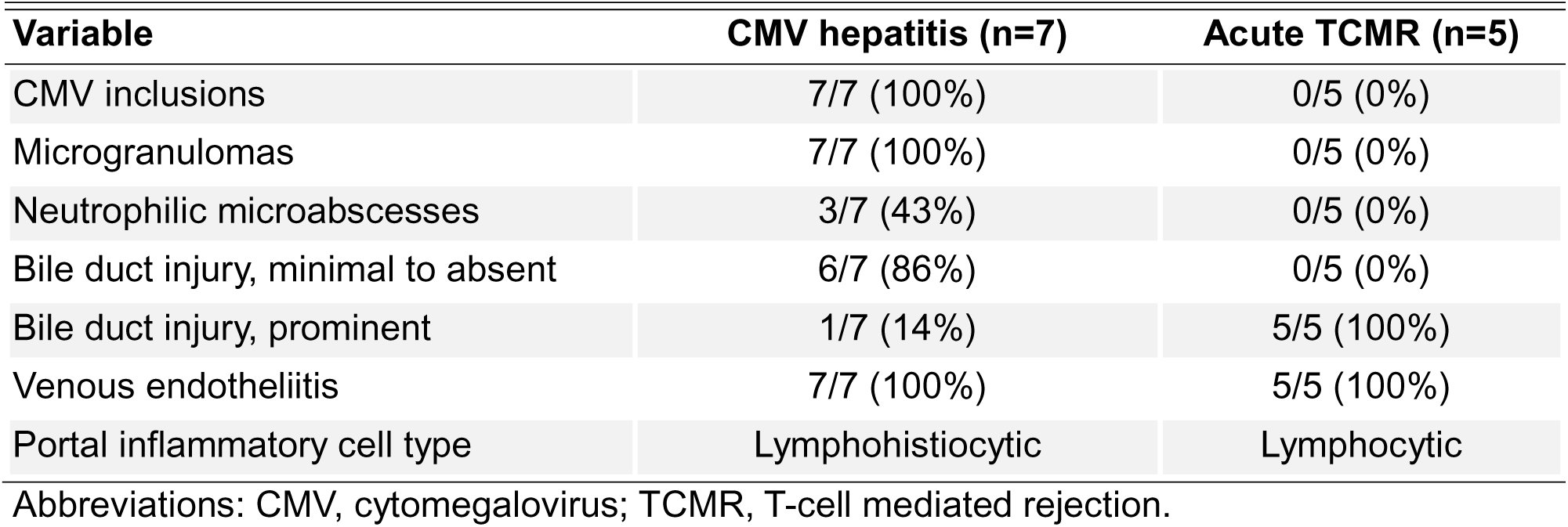
Histopathologic features.

### Diagnostic discrimination

To quantify the discriminatory value of histologic features, statistical analyses were performed to calculate the sensitivity and specificity of each feature. CMV inclusions showed sensitivity 100% (95% CI 59.0–100.0), specificity 100% (95% CI 47.8–100.0), p=0.0013. Microgranulomas also showed sensitivity 100% (95% CI 59.0–100.0) and specificity 100% (95% CI 47.8–100.0), p=0.0013. Bile duct injury minimal to absent showed sensitivity 86% (95% CI 42.1–99.6) and specificity 100% (95% CI 47.8–100.0), p=0.015. Neutrophilic microabscesses showed sensitivity 43% (95% CI 9.9–81.6) and specificity 100.0%, p=0.205. Results are shown in **Table 3**.

**Table 3.** Diagnostic performance of histologic features.

| **Histologic feature** | **CMV hepatitis** | **Acute TCMR** | **Sensitivity, % (95% CI)** | **Specificity, % (95% CI)** | **p value** |
| --- | --- | --- | --- | --- | --- |
| CMV inclusions | 7/7 | 0/5 | 100.0% (59.0–100.0) | 100.0% (47.8–100.0) | 0.0013 |
| Microgranulomas, portal and/or lobular | 7/7 | 0/5 | 100.0% (59.0–100.0) | 100.0% (47.8–100.0) | 0.0013 |
| Bile duct injury, minimal to absent | 6/7 | 0/5 | 85.7% (42.1–99.6) | 100.0% (47.8–100.0) | 0.015 |
| Neutrophilic microabscesses | 3/7 | 0/5 | 42.9% (9.9–81.6) | 100.0% (47.8–100.0) | 0.205 |

Abbreviations: CMV, cytomegalovirus; TCMR, T-cell mediated rejection.

### Management and follow-up

Antiviral therapy was given in 9 of 10 CMV patients (**Table 4**). In the CMV cohort, acute TCMR at the index biopsy was present in one patient and indeterminate in four patients, and five had no acute rejection, with one of the latter diagnosed as early chronic rejection. Anti-rejection therapy was administered in one of the four indeterminate cases and in two of the five without acute rejection, whereas the single patient with a histologic diagnosis of concomitant TCMR did not receive such therapy. All acute TCMR controls received anti-rejection therapy.

**Table 4.** Treatments and outcomes.

| **Variable** | **CMV hepatitis (n=10)** | **Acute TCMR (n=5)** |
| --- | --- | --- |
| Antiviral therapy | 9/10 (90%) | 0/5 (0%) |
| Anti-rejection therapy | 3/10 (30%) | 5/5 (100%) |
| CMV recurrence/viremia | 4/10 (40%) | 0/5 (0%) |
| Progression to late chronic rejection | 2/10 (20%) | 0/5 (0%) |

Abbreviations: CMV, cytomegalovirus; TCMR, T-cell mediated rejection.

During follow11up, recurrent CMV viremia occurred in four patients in the CMV group; among these, one subsequently progressed to late chronic rejection and another had an indeterminate result on a later biopsy. In addition, one patient without documented CMV recurrence progressed to late chronic rejection. No CMV viremia was observed in the acute TCMR group.

### Representative Case

Among the CMV group, one illustrative example was a man in his 60s with CMV D+/R− serostatus who underwent liver transplantation for autoimmune hepatitis and metabolic-associated steatohepatitis. Within the first year after transplantation, he presented with malaise and elevated liver enzymes (ALT 96 U/L, AST 106 U/L, ALP 243 U/L). Liver biopsy revealed moderate inflammatory infiltrates composed of lymphocytes, eosinophils, neutrophils, and macrophages with bile duct injury and interface activity (**Figure 1A**), initially suggestive of acute T-cell–mediated rejection. Lobular activity with poorly formed non-necrotizing granulomas was also present. On careful examination, a single epithelial cell with intranuclear and intracytoplasmic inclusions characteristic of CMV infection was identified (**Figures 1B-C**), establishing the diagnosis of CMV hepatitis. Plasma CMV PCR was positive (914 IU/mL). Special stains (acid-fast bacilli, Fite, Grocott, and periodic acid-Schiff with diastase) were negative for other infectious organisms. Due to clinical uncertainty regarding concurrent rejection, intravenous steroids and intravenous ganciclovir were administered, resolving CMV viremia and normalizing liver enzymes within one month; subsequent low-level recurrent viremia responded to valganciclovir. Immunosuppression was gradually minimized, with stable graft function thereafter.

**Figure 1.**
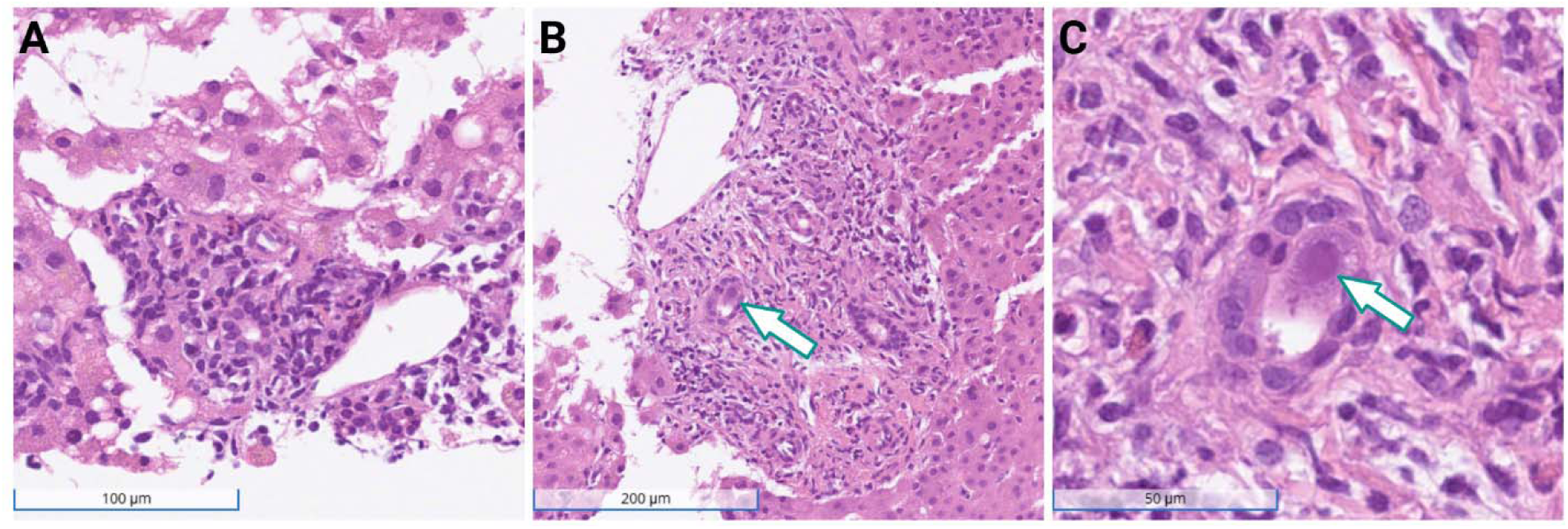
Cytomegalovirus hepatitis in a liver allograft biopsy. H&E shows (A) portal mixed inflammation with bile duct injury and interface activity, mimicking T-cell–mediated rejection. (B-C) A single epithelial cell with intranuclear and intracytoplasmic CMV inclusions (arrow), shown at lower (B) and higher (C) magnification.

## Discussion

In this single-center cohort, CMV hepatitis showed a characteristic pattern that helps separate it from acute TCMR during routine sign-out: portal-predominant inflammation with a histiocyte-rich component, lobular microgranulomas, and occasional neutrophilic microabscesses in the setting of little or no bile duct injury.^7, 10^ These observations align with recent series emphasizing that CMV can convincingly resemble rejection at low power.^8^

Venous endotheliitis occurred in both groups and was therefore not discriminatory in our material, consistent with prior work.^6, 8^ Sinusoidal endotheliitis has been proposed as a supplemental marker for T TCMR and correlates with the endotheliitis component of the Banff rejection activity index, but it is not specific when considered in isolation in the post-transplant liver.^11^ In practice, endotheliitis should be weighed with the character of the portal infiltrate, the degree of bile duct injury, and the lobular background, with deeper levels and CMV immunohistochemistry or plasma PCR when morphology is equivocal.^7, 8, 10^

Our findings should be interpreted alongside recent Banff Liver Group work that distinguishes early from late TCMR.^12, 13^ Late-onset cases are enriched for interface hepatitis and central perivenulitis, with a lymphocyte predominant infiltrate and fibrosis, and show less conspicuous bile duct-centered injury and portal subendothelial inflammation than early-onset disease. Central perivenulitis, a zone 3 perivenular inflammatory pattern, is distinct from central venulitis and is a recognized feature in late post-transplant liver allograft biopsies.^12, 14^ Because our control group consisted of acute T-cell–mediated rejection, the present data primarily address separation of CMV hepatitis from the early phenotype. Validation in cohorts that include late-onset rejection is warranted.

We observed discordance between pathology reports and therapy. Some patients without definite acute TCMR received anti-rejection treatment, while one diagnosed with acute TCMR did not. Together with recurrent CMV viremia and subsequent chronic rejection in a minority of patients, these patterns highlight the need to integrate pathology with contemporaneous virology when treatment decisions are made.^1^

CMV and TCMR are not mutually exclusive. CMV can potentiate or trigger TCMR through cross-reactivity, immune activation, and endothelial stimulation, while alloimmune activation can, in turn, favor CMV replication; together these observations indicate a two-way interaction.^15–18^ There is also evidence that controlling CMV may lessen rejection activity in selected settings. In liver allografts, observational data link antiviral prophylaxis to fewer episodes of biopsy-proven acute rejection.^19^ Moreover, a recent pathology series reported that a subset of CMV hepatitis cases that mimicked acute cellular rejection improved with antiviral therapy alone.^8^ In other solid organ cohorts, intensified anti-CMV strategies have been associated with fewer rejection events, including reduced acute rejection after heart transplantation and lower early subclinical rejection with prophylaxis compared with preemptive therapy after kidney transplantation.^20–22^ In a representative case from our cohort, once a viral inclusion was identified, antiviral therapy was initiated, and liver tests improved without escalation of immunosuppression.

The relationship between CMV and chronic rejection remains unsettled. In explant and biopsy series from the era before routine ganciclovir prophylaxis, persistent CMV genomes were localized to structures central to chronic rejection pathogenesis, with strong signals in residual bile ducts and moderate signals in vascular endothelium in grafts lost to chronic rejection; all 10 patients in one series had post-transplant CMV infection and 9 of 9 available explants were CMV-DNA positive.^23^ In a prospective cohort of 33 patients undergoing 57 transplants, the combination of donor CMV-seronegative with recipient seropositive status and CMV infection persisting more than 30 days was associated with increased relative risk of chronic rejection, whereas the mere presence of infection, symptomatic disease, serum or urine PCR positivity, and peak or cumulative viral load were not predictive.^24^ Mechanistic reviews describe plausible pathways by which CMV could accelerate ductopenic and vasculopathic injury, including upregulation of endothelial adhesion molecules and HLA expression, skewing toward humoral responses, and profibrotic signaling that promotes biliary epithelial injury and graft arteriosclerosis.^25^ Contemporary overviews continue to recognize links between CMV and long-term graft injury while acknowledging variability in effect size across studies and eras.^5, 17^ By contrast, in a 12-year single-center series of 1,146 liver transplants in which biopsy-proven CMV hepatitis occurred in 2.1%, no long-term correlation with chronic rejection was observed despite associations with D+/R-status and OKT3 treatment.^26^ In our cohort, 2 of 10 recipients with CMV infection later developed late chronic rejection, which is compatible with prior reports, although causality cannot be established in our small series.

The strengths of this study include consistent biopsy handling at a single institution, review by a board-certified pathologist with expertise in medical liver pathology, and the use of predefined histologic variables from Banff schema, which improves reproducibility. Linking histology to treatment and short-term outcomes further highlights the clinical impact of initial classification.

Limitations include the retrospective design, single-observer non-blinded re-review, small cohort, and incomplete slide availability in some CMV patients. Accordingly, the estimated sensitivity and specificity of individual histologic features should be interpreted as exploratory rather than confirmatory. Sampling variability is a concern because inclusions may be very focal, and CMV immunohistochemistry was not consistently performed throughout the study period. The control group included only patients with moderate to severe acute TCMR who were CMV PCR negative, which clarified the contrast with CMV hepatitis but excluded the rare situation of concurrent CMV infection and clinically significant rejection. In addition, donor11specific antibody testing and C4d immunohistochemistry were not available, precluding analysis of antibody11mediated mechanisms in patients who progressed to chronic rejection, an issue highlighted in recent Banff discussions.^12^ All available slides were re-reviewed by a single liver pathologist; however, because this study was designed to distinguish CMV hepatitis from acute TCMR, we prespecified histologic variables accordingly and did not apply standardized scoring of interface hepatitis or central perivenulitis. As a result, features typical of late-onset TCMR were not analyzed systematically. Larger prospective studies should include routine DSA testing, C4d staining, and standardized masked scoring of interface hepatitis and central perivenulitis.

In summary, lobular microgranulomas and minimal to absent bile duct injury were the most informative histologic clues for favoring CMV hepatitis over TCMR in this cohort. When interpreted alongside a portal lymphohistiocytic infiltrate, and paired with targeted inclusion searches and CMV PCR, this pattern can reduce unnecessary intensification of immunosuppression and enable timely antiviral therapy, with potential to improve graft outcomes.

## Ethical Approval

This study was approved by the Institutional Review Board at the Hospital of the University of Pennsylvania, meeting exemption criteria (category 4) with a HIPAA waiver. This study was conducted in accordance with the Declaration of Helsinki 1975.

## Informed Consent

In accordance with institutional policy, the hospital obtains general consent at the time of care that permits secondary research use of de-identified clinical data and archived pathology material; therefore, the Institutional Review Board waived the requirement for study-specific informed consent.

## Declaration of Conflicting Interests

None declared.

## Funding

This study did not receive any funding.

## Data Availability Statement

All data produced in the present study are available upon reasonable request to the authors.

## Notes

### Competing Interest Statement

The authors have declared no competing interest.

### Author Declarations

This study was approved by the Institutional Review Board at the Hospital of the University of Pennsylvania, meeting exemption criteria (category 4) with a HIPAA waiver.

### Summary of Updates

Tables 2-4 have been added.

